# Microbubbles for improved nucleic acid extraction in wastewater samples for viral RNA detection

**DOI:** 10.1101/2022.07.10.22277342

**Authors:** Casey Wegner, Jon Roussey, Maureen Carey, Brittany Macintyre, Tiffany Snow, Douglas Sieglaff, Aline Bridi, Shauntina Battiste, Carolina B. Livi, Brandon McNaughton

**Author notes:** **Corresponding Author:** Casey Wegner –.

## Abstract

The use of wastewater-based epidemiology has increased in recent years due to the publication of COVID-19 online trackers and the focus of the media on the pandemic. Yet the analysis of viromes in wastewater has been widely applied for several decades in conjunction with traditional chemical analysis approaches. However, even though real time quantitative polymerase chain reaction (RT-qPCR) based molecular detection methods are now mainstream in large and small labs alike, wastewater sampling and nucleic acid extraction procedures are not yet standardized or optimized to enable routine and robust analysis and results interpretation. Here, we employ a flotation-based nucleic acid extraction method using microbubbles that allows for simple direct collection and lysis of total wastewater samples without the requirement for pasteurization or filtration of solid components prior to analysis. An additional advantage discovered during testing was reduced sample input needs while maintaining sensitivity compared to precipitation and ultrafiltration-based methods. Microbubbles designed to bind nucleic acids enable convenient workflows, fast extraction, and concentration and purification of RNA and DNA that is compatible with downstream genomic analyses.

**SUMMARY:** Microbubble-based capture of nucleic acids from raw (unpasteurized) and unfiltered (containing solids) wastewater with subsequent elution offers several advantages over existing methods. Using microbubbles, the required sample input and protocol duration are reduced while sensitivity of downstream genomic analysis is increased.

## INTRODUCTION

### Importance of Wastewater Testing

One of the most challenging problems during the SARS-CoV-2 pandemic has been monitoring the state of the outbreak in real time (Wu et al., 2020; Singh and Yi, 2021). This was especially true when SARS-CoV-2 first emerged as there were not enough diagnostic tests to sample entire populations, resulting in a need for pooling and a massive ramp up in assay kit manufacturing. The problem has been further compounded by resistance to testing among some individuals. The development of at-home tests has added convenience for individuals but made monitoring numbers of cases even more challenging. This has led to expanded use of established wastewater epidemiology methods for SARS-CoV-2 monitoring at the population level (Hart and Halden, 2020; Peccia et al., 2020; Daigle et al., 2022; Qiu et al., 2022). Several college campuses have adopted a localized testing strategy for evidence-based decision making in real time during the pandemic (Karthikeyan et al., 2021; Harris-Lovett et al., 2021; Anderson-Coughlin et al., 2022). The CDC has incorporated many US wastewater sites into its monitoring program and many public online dashboards are updated frequently.

### Status quo and challenges of wastewater testing

While molecular detection for infectious organisms in wastewater has been in use for several decades, it is an inherently complex sample type with high levels of particulates, organic molecules and diverse biologically derived materials (Ye et al., 2016). In fact, human viruses of interest end up at wastewater plants which collect feces, urine and sputum along with other human cells and can be detected using microbiological, proteomic, or genomic assays. Wastewater-based epidemiology has been used for several decades to monitor pathogens including viruses such as influenza and poliovirus, the causative agents of flu and polio (Heijnen and Medema, 2011; Brouwer et al., 2018). The complexity and instability of the sample and associated health risks has led to limitations of the methods available to both extract and analyze nucleic acids including RNA. Many new protocols and methods continue to use longstanding approaches for extraction, such as precipitation, filtration and magnetic separation.

Several studies highlight the challenges with current wastewater testing methodologies (Ahmed et al., 2020; Sharkey et al., 2021; Pecson et al., 2021) and mention a need for increased frequency of sampling and a decrease in the number of steps and elapsed time between collection and analysis. Here we describe a method relying on lysis of raw wastewater samples that include solids followed by enrichment of nucleic acids using functionalized microbubbles that addresses several limitations of currently implemented strategies.

### Microbubbles as a viable solution

Tracking the virome in wastewater involves extraction of nucleic acids followed by detection of DNA or RNA with sequences derived from specific microorganisms. These methods have found use in many wastewater workflows but have limitations that restrict broader adoption and implementation of monitoring. For example, precipitation requires an overnight protocol step and filtration struggles with clogging due to the high number of particulates. Both methods require large volumes of wastewater input samples. A need for increased frequency of sampling, reduced protocol steps and improved time to results has been identified by the testing community as referenced above. Here we demonstrate the use of microbubbles for nucleic acid extraction, allowing for simple, direct collection and lysis of total wastewater samples without requirement for pasteurization or filtration of solid components prior to nucleic acid purification and subsequent analysis.

## PROTOCOL

### 3.0 mL Wastewater Akadeum RNA Extraction Procedure

Before you begin:

- Prepare all necessary supplies nuclease-free.
- Perform all steps at room temperature.
- All centrifugation steps should be performed in swinging bucket centrifuges.
- Preheat a water bath to 60 ° C and warm Buffer ES (ES) to 60 ° C.

1.0 Sample Lysis
  1.1 Add 3.0 mL of wastewater to 15 mL centrifuge tube.
  1.2 To samples < 3.0 mL add 1X PBS to a total volume of 3.0 mL. *Add any controls or engineered metagenomic spike-ins at this stage*. Mix well.
  1.3 Add 5.0 mL of Buffer LS; mix by vortexing for 5 seconds.
  1.4 Add 150 µL Reagent A; mix by vortexing for 5 seconds.
  1.5 Incubate at 60 ° C for 10 minutes.

2.0 Nucleic Acid Extraction and Binding
  2.1 Add 5.0 mL 100% EtOH and mix by vortexing for 5 seconds.
  2.2 Add 1.0 mL Akadeum RNA Extraction Microbubbles; mix by end-over-end rotation at ≥ 20 rpm for five (5) minutes.
  2.3 Centrifuge at 3000 g for 30 seconds or allow bubbles to rise by flotation for five (5) minutes.
  2.4 Being very careful not to disturb bubble layer, aspirate and discard subnatant and pellet with a 9” Pasteur pipette; there should be < 0.5mL of subnatant below the bubble layer. Note: user will need to complete several (∼4-5) transfers of the subnatant volume from the tube to waste with the 9” Pasteur pipette.
  2.5 Add 2.5 mL 80% EtOH and vortex for three (3) seconds.
  2.6 Centrifuge at 3000 g for 20 seconds or allow bubbles to rise by flotation for five (5) minutes.
  2.7 Aspirate and discard subnatant below the bubble layer using a 9” Pasteur pipette.

3.0 Nucleic Acid Elution and Concentration
  3.1 Add 400 µL 60 ° C Buffer ES to microbubbles; incubate sample for two (2) minutes at room temperature.
  3.2 Vortex for 15 seconds. Ensure all microbubbles are in solution before proceeding.
  3.3 Centrifuge at 3000 g for 20 seconds or allow bubbles to rise by flotation for five (5) minutes.
  3.4 Add 550 µL Buffer LS and 525 µL 100% EtOH to a new 2.0 mL tube.
  3.5 Transfer 425 µL subnatant into the 2.0 mL tube containing the Buffer LS-EtOH solution. Mix by pipetting. Note: Residual ethanol disassociates from microbubbles following addition of buffer, increasing total volume beneath the microbubbles to > 400 µL.
  3.6 Transfer 750 µL of the sample to a spin column (column placed in a 2.0-mL collection tube). Centrifuge for 15 seconds at ≥ 8000 g.
  3.7 Discard flow-through; repeat step 3.6 with remaining sample as many times as needed until the entire sample has been passed through the spin column. Discard flow-through.
  3.8 Add 500 µL 80% EtOH to the spin column. Centrifuge for 2 minutes at ≥ 8000 g.
  3.9 Place the spin column in a new 1.5 mL collection tube. Add 50 µL 60 °C Buffer ES and incubate sample fortwo (2) minutes at room temperature.
  3.10 Centrifuge for one (1) minute at ≥ 8000 g.
  3.11 Collect filtrate for downstream use: use immediately or store at −20 °C for up to one week or −80 °C for over one week.

### 10 mL Wastewater Akadeum RNA Extraction Microbubbles Procedure

Before you begin:

- Prepare all necessary supplies nuclease-free.
- Perform all steps at room temperature.
- All centrifugation steps should be performed in swinging bucket centrifuges.
- Preheat a water bath to 60 ° C and warm Buffer ES (ES) to 60 ° C.
- Optional: The Procedure can leverage a custom microbubble drain tube to eliminate the need for Pasteur pipetting steps.

1.0 Sample Lysis
  1.1 Add 10.0 mL of wastewater to 50 mL centrifuge tube.
  1.2 To samples < 10.0 mL add 1X PBS to a total volume of 10.0 mL. *Add any controls or engineered metagenomic spike-ins at this stage*. Mix well.
  1.3 Add 17 mL of Buffer LS; mix by vortexing for five (5) seconds.
  1.4 Add 475 µL Reagent A; mix by vortexing for five (5) seconds.
  1.5 Incubate at 60 ° C for 10 minutes.

2.0 Nucleic Acid Extraction and Binding
  2.1 Add 17 mL 100% EtOH and mix by vortexing for five (5) seconds.
  2.2 Add 2.0 mL Akadeum RNA Extraction Microbubbles; mix by end-over-end rotation at ≥ 20 rpm for five (5) minutes.
  2.3 Centrifuge at 3000 g for 30 seconds or allow bubbles to rise by flotation for five (5) minutes.
  2.4 Being very careful not to disturb bubble layer, aspirate and discard subnatant and pellet with a 9” Pasteur pipette; there should be < 0.5mL of subnatant below the bubble layer. Note: user will need to complete several (∼15) transfers of the subnatant volume from the tube to waste with the 9” Pasteur pipette.
  2.5 Add 5.0 mL 80% EtOH and vortex for three (3) seconds
  2.6 2.6 Centrifuge at 3000 g for 20 seconds or allow bubbles to rise by flotation for five (5) minutes.
  2.7 Aspirate and discard subnatant below the bubble layer using 9” Pasteur pipette.

3.0 Nucleic Acid Elution and Concentration
  3.1 Add 975 µL 60 ° C Buffer ES to microbubbles; incubate sample for two (2) minutes at room temperature.
  3.2 Vortex for 15 seconds. Ensure all microbubbles are in solution before proceeding.
  3.3 Centrifuge at 3000 g for 20 seconds or allow bubbles to rise by flotation for five (5) minutes.
  3.4 Add 1275 µL Buffer LS and 1225 µL 100% EtOH to a new 5.0 mL tube.
  3.5 Transfer 1000 µL subnatant into the 5.0 mL tube containing the Buffer LS-EtOH solution. Mix by pipetting. Note: Residual ethanol disassociates from microbubbles following addition of buffer, increasing total volume beneath the microbubbles to >975 µL.
  3.6 Transfer 750 µL of the sample to a spin column (column placed in a 2.0-mL collection tube). Centrifuge for 15 seconds at ≥ 8000 g.
  3.7 Discard flow-through; repeat step 3.6 with remaining sample as many times as needed until the entire sample has been passed through the spin column. Discard flow-through.
  3.8 Add 500 µL 80% EtOH to the spin column. Centrifuge for 2 minutes at ≥ 8000 g.
  3.9 Place the spin column in a new 1.5 mL collection tube. Add 50 µL 60 °C Buffer ES and incubate sample for two (2) minutes at room temperature.
  3.10 Centrifuge for one (1) minute at ≥8000 g.
  3.11 Collect filtrate for downstream use: use immediately or store at −20 °C for up to a week or −80 °C for over a week.

## REPRESENTATIVE RESULTS

RNA and DNA recovery was compared following two different microbubble extraction protocols from 3 mL of untouched wastewater. Protocol A was a protocol that was developed for microbubble nucleic acid extraction from saliva samples. Protocol B represents a protocol that was optimized for maximizing nucleic acid extractions from wastewater samples. The protocol optimizations that resulted in improved RNA and DNA recovery included reformulation of buffers LS and ES, switching to a different nucleic acid spin column manufacturer, and utilizing heat during elution. RNA and DNA recoveries from wastewater for Protocols A and B were measured in triplicate by Qubit fluorometric quantification and corresponding high-sensitivity RNA and DNA kits. Protocol B is shown in the Protocol section.

Wastewater influent samples were evaluated for the presence of SARS-CoV2 following RNA extraction with microbubbles. Positive wastewater samples—previously characterized for SARS-CoV-2 presence following PEG precipitation based nucleic acid extraction—were provided by the University of Michigan. The samples were collected from five (5) locations in Michigan (Ann Arbor, Flint, Jackson, Tecumseh and Ypsilanti). For each sample, two 3 mL extractions were performed. Extracted nucleic acid eluates were run in duplicate with Perkin-Elmer SARS-CoV2 RT-PCR kit on the ABI 7900 384 well Real-time PCR instrument. The results demonstrate that detection concordance was observed between a conventional PEG precipitation RNA extraction method (40 mL sample input) and microbubble RNA extraction with RT-qPCR analysis (3 mL wastewater sample input).

SARS-CoV-2 RNA (ATCC strain VR-3447), 1E4 copies/mL, was spiked into wastewater influent that did not contain detectable levels of SARS-CoV-2 RNA. Microbubble extractions, from either 10 mL or 3 mL of SARS-CoV-2 spiked wastewater volumes, were performed with nucleic acid microbubbles. Extracted RNA was evaluated with the Perkin Elmer RT-PCR for SARS-CoV-2 Kit on the ABI 7900 Real time PCR instrument. The wastewater influent catchments were collected at the Flint City Wastewater Plant, Flint, Michigan. Significantly increased (two-tailed t-test) levels of SARS-CoV2 RNA from wastewater was detected following microbubble nucleic acid extraction from the 10 mL input samples as compared to the 3 mL input samples.

The next set of experiments focus on establishing the flotation-based methods in lieu of currently adopted wastewater sample processing methods for SARS-CoV-2 testing based on RT-qPCR. Table 3 describes the methods compared to the flotation-based method optimized for wastewater. We include a summary of materials and methods as they differ slightly from the finalized protocol presented.

**Table 1.** Microbubble nucleic acid extraction protocol optimized for wastewater samples.

*Nucleic Acid Extraction Comparison from Wastewater*
| **Protocols** | **RNA (ng/$\mu\text{L}$)** | **DNA (ng/$\mu\text{L}$)** |
| --- | --- | --- |
| *A* | 6.84 ± 0.64 | 18.7 ± 1.86 |
| *B* | 11.90 ± 0.84 | 27.23 ± 2.89 |
| *% Change* | +74.0 % | +45.6 % |

**Table 2.** Microbubble RNA extraction concordance with PEG precipitation and high-sensitivity molecular analysis.

| Sample ID | PEG Precipitation (40 mL) | Microbubble Extraction (3 mL) |
| --- | --- | --- |
|  | Previously characterized | qPCR |
|  | Target Detected<br>(N1 and/or N2 gene) | Target Detected<br>(N1 gene) |
| 26AA220529I | Yes | Yes |
| 26FL220528I | Yes | Yes |
| 26FL220529I | Yes | Yes |
| 26FL220530I | Yes | Yes |
| 26JS220629I | Yes | Yes |
| 26JS220630I | Yes | Yes |
| 26TM220628I | Yes | Yes |
| 26TM220529I | Yes | Yes |
| 26TM220630I | Yes | Yes |
| 126YC220628I | Yes | Yes |
| Negative Control<br>(PBS) | N/A | No |

**Table 3.** Wastewater Processing Method Comparison.

| Material/ Process | Akadeum | UF + ABS | PEG/NaCl + ABS |
| --- | --- | --- | --- |
| WAW volume | 3 or 10 mL | 45 mL | 45 mL |
| Spike-in incubation | 4°C for 90 min | 4°C for 90 min | 4°C for 90 min |
| Pasteurization | 60 or 75°C for 90 min* | 75°C for 90 min | 60°C for 90 min |
| Removal of TSS | N/A | 0.22 µM vacuum filtration device | 3,000 x g 15 minutes (4°C) |
| Concentration | Via microbubble Binding | 10 kDa NMWL ultrafiltration device | 10% PEG/ 0.3 M NaCl (overnight @ 4°C) |
| RNA Extraction | Microbubbles + clean-up w/ Absolutely RNA Binding Cup | Absolutely RNA Miniprep Kit | Absolutely RNA Miniprep Kit |
| RNA elution reagents | Elution Buffer | Elution Buffer | Elution Buffer |
| RNA elution volume | 100 µL | 100 µL | 100 µL |
| WAW (mL) Equivalents/ qRT-PCR | 3 mL x (0.005 mL/0.1 mL) = 0.15 mL<br>10 mL x (0.005 mL/0.1 mL) = 0.50 mL | 45 mL x (0.005 mL/0.1 mL) = 2.25 mL | 45 mL x (0.005 mL/0.1 mL) = 2.25 mL |
| Estimated Elapsed time from WAW sample to RNA | 1.5 hours (excludes pasteurization*) | 3 hours | 17 hours (includes overnight precipitation at 4°C) |

\*Note, pasteurization is unnecessary with the Akadeum microbubble protocol as a chaotropic lysis buffer is added to raw influent as the first step. TSS = Total Suspended Solids; NMWL = Nominal molecular weight limit; Elution Buffer = 10 mM Tris-HCl (pH 7.5) and 0.1 mM EDTA

For the results presented in Figure 2, wastewater influent grab samples were obtained from the Walnut Creek Wastewater Treatment Plant (Austin, TX), and transported to the laboratory for analysis. Prior to testing, all wastewater was mixed to bring settled solids back into solution, then aliquoted for processing. Wastewater spiked with viruses (SARS-CoV-2, ATCC, VR-1986HK; Human coronavirus 229E, ATCC, VR-740) was tested, as well non-spiked wastewater (assesses endogenous SARS-CoV-2). Pepper Mild Mottle Virus (PMMoV) was also analyzed in all samples as an indication of population fecal load. While not required, wastewater samples were pasteurized at 60oC or 75°C prior to wastewater concentration methodology.

**Figure 1.**
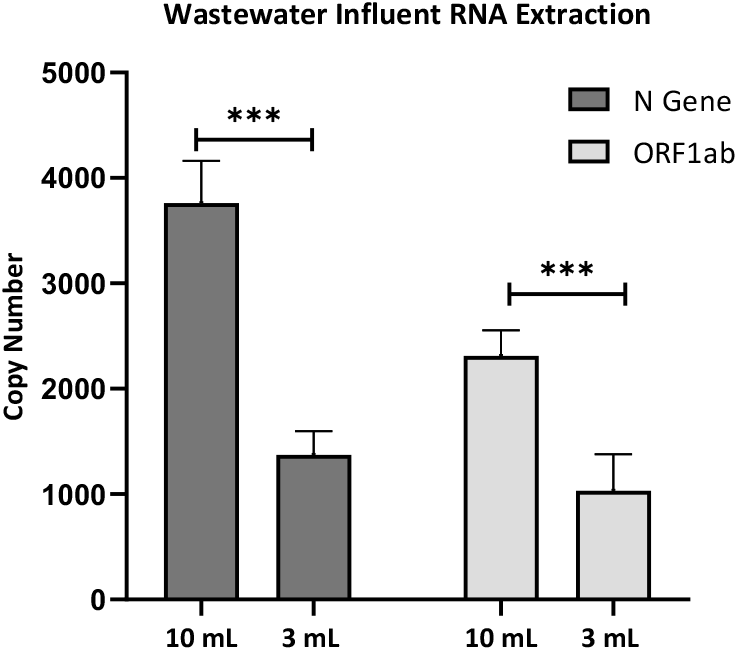
Microbubble RNA extraction from larger wastewater volumes enhances RT-qPCR sensitivity.

**Figure 2.**
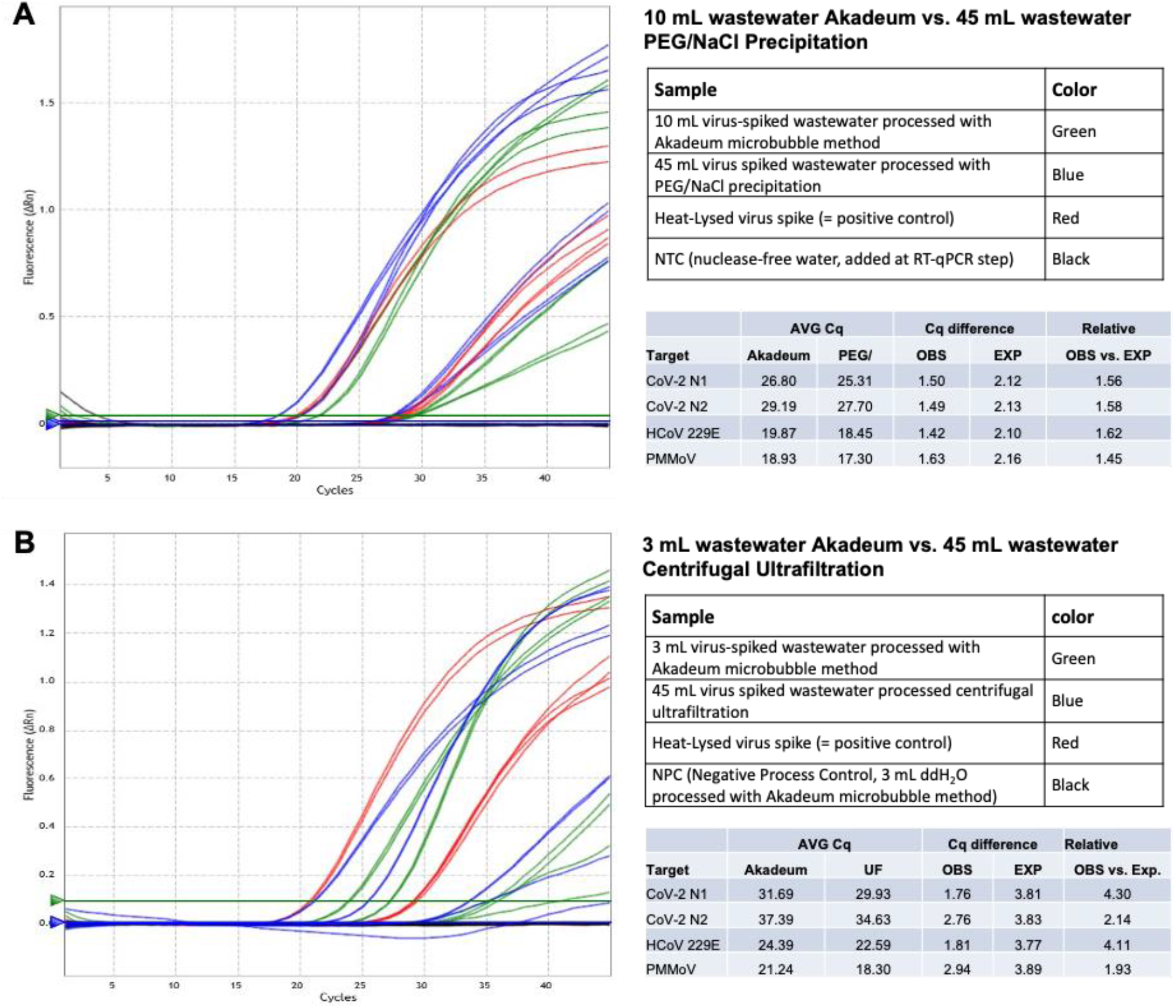
qPCR results comparison. A: Akadeum microbubble and PEG/NaCl Precipitation and B: Akadeum microbubble and Centrifugal ultrafiltration. Color legend in first table. Cq values and Cq difference calculated as: OBS = AVG Akadeum Cq – AVG Comparator Cq; EXP = LOG(volumetric ratio, target slope base log); Volumetric Sensitivity Equivalent (VSE) = 45 mL / (target slope base log(EXP-OBS))

Two commonly practiced methods for wastewater concentration and viral processing were employed as a comparator to the described Akadeum microbubble buoyancy activated protocol, centrifugal ultrafiltration (UF) and PEG/NaCl (PEG) precipitation, both processing 45 mL of wastewater. UF and PEG require the removal of larger suspended solids prior to wastewater concentration; vacuum filtration or centrifugation for the UF and PEG protocols, respectively (Pecson et al., 2021) was used for solids removal. Akadeum microbubbles were used to process the entire wastewater sample (i.e., included both suspended solids and aqueous phase).

Several conditions using functionalized microbubbles to bind to nucleic acids developed for RNA extraction of SARS-CoV-2 with detection by RT-qPCR were tested to evaluate use of the microbubbles for wastewater testing. 3.0 and 10 mL microbubble RNA extraction in wastewater were carried out according the Protocol. The eluted RNA was analyzed by qRT-PCR using CDC SARS-CoV-2 N1 and N2 designed primer/probes, and custom designed primer/probes for HCoV 229E and PMMoV, using Agilent Brilliant III Ultra-Fast qRT-PCR Master Mix (Agilent Technologies) and Agilent AriaMx real-time PCR System. Quantitative RT-PCR gene target slopes were derived from 3 independent viral spiked wastewater samples. The slopes were then used to compare expected Cq differences between methods based upon wastewater volumes processed to the observed Cq differences between methods. For example, with a gene target slope of −3.32 (base log = 2) the expected qRT-PCR cycle difference between processing 45 and 3.0 mL of wastewater = −3.91 (i.e., log2(3/45)). If the observed qRT-PCR cycle difference between 45 mL (UF) and 3 mL (Akadeum) = −2.01, the 3 mL (Akadeum) method displayed 3.72 higher observed sensitivity relative to expected, based upon wastewater volumes processed (i.e., 2((−2.01)-(−3.91))). This value was then used to derive Volumetric Sensitivity Equivalent (VSE) = 45 mL/ 3.72 = 12.1 mL (i.e., the flotation-based method requires 12.1 mL to achieve equivalent sensitivity to the comparator method processing 45 mL).

Figure 2 shows amplification plots and the number of cycles to reach established thresholds from RT-qPCR measurements obtained from RNA samples isolated using the methods described in Table 3.

## DISCUSSION

Microbubbles have been used for many applications, including cell isolations—more commonly known as Buoyancy Activated Cell Sorting (Akadeum Life Sciences, 2020; McNaughton et al., 2018; Jeon, D. & McNeel, D., 2022; Brooks et al., 2021), dead cell removal (Akadeum Life Sciences, 2022; McNaughton et al., 2018), cell therapy (Snow et al., 2022) and nucleic acid extractions from saliva (Akadeum Life Sciences, 2021). As detailed above, we have demonstrated a new protocol and application for microbubbles: viral nucleic acid extraction from wastewater using microbubbles.

The COVID-19 pandemic highlighted how biomedical molecular testing technologies have advanced and can be applied to urgent global needs (Solo-Gabriele et al., 2022). Implementing testing programs also exposed limitations in sample collection, handling, and processing as well as ability to implement longitudinal monitoring at scale. The ability to pool samples assessing viral burden in small groups or communities for decision making instead of relying exclusively on individual testing removed some of the pressure in terms of number of samples to be run. This requires the ability to process higher volumes while maintaining sensitivity for a single positive input to still be detected.

In fact, when combined with RNA binding columns and RT-qPCR, this workflow requires significantly less sample volume (as shown in Figure 2) and has equivalent to increased sensitivity compared to commonly practiced wastewater processing methods on a volumetric level. One of the strengths of floatation-based separation is that it can be scaled-up in volume quite dramatically due to the uniform force of buoyancy that drives the separation of the microbubbles away from the bulk wastewater. As a result, it is straightforward for the sample volume to be scaled up if total nucleic acid yields need to be increased. Alternatively, less sample as compared to standard methods, can be used in the microbubble workflow (*i*.*e*. it is possible to lower limits of detection for post treatment water analysis without increasing sample volume inputs).

Another advantage is an improved or simplified workflow. In particular, the described workflow enables assessment of wastewater without the need for pre-processing, such as heat treatment or the removal of solids from influent samples. Due to the uniformity of separating force, complex sample matrices, such as wastewater, can be readily processed. Furthermore, by use of a nucleic acid extraction method, pasteurization (i.e., direct addition of lysis buffer to the raw wastewater) is not required. This as well as biosafety issues with sample handling are addressed using this lysis followed by nucleic acid binding microbubble approach.

It is clear the need for monitoring of human viruses in the build environment will continue and likely expand as climate change and a globalized supply chain facilitates long range disease spread. This will include using wastewater, air filters and surface swabs in addition to testing individuals for SARS-CoV-2 (Zeller et al., 2021; Solo-Gabriele et al., 2022) as well as other infectious viruses of interest (Botti-Lodovico et al., 2021). With vaccinations unable to fully contain COVID-19 pandemic (Keehner et al., 2021) and universal mask wearing for long periods of time impractical and difficult to enforce, frequent wastewater monitoring using RT-qPCR and other genomics measurements can detect and enable health care alerts to prepare agencies and providers in affected areas. Floatation-based nucleic acid extraction from wastewater provides a solution that addresses inactivating viral particles, stabilizing analyte and minimizing equipment requirements for sample collection and processing.

## Data Availability

All data produced in the present work are contained in the manuscript

## ACKNOWLEDGMENTS

The authors would like to thank the University of Michigan and Ivan Tapia and the Walnut Creek Wastewater Treatment Plant team for providing wastewater samples.

